# Phe2vec: Automated Disease Phenotyping based on Unsupervised Embeddings from Electronic Health Records

**DOI:** 10.1101/2020.11.14.20231894

**Authors:** Jessica K. De Freitas, Kipp W. Johnson, Eddye Golden, Girish N. Nadkarni, Joel T. Dudley, Erwin P. Bottinger, Benjamin S. Glicksberg, Riccardo Miotto

**Affiliations:** Hasso Plattner Institute for Digital Health at Mount Sinai, Icahn School of Medicine at Mount Sinai, 1 Gustave L. Levy Pl, New York, NY 10029, USA; Department of Genetics and Genomic Sciences, Icahn School of Medicine at Mount Sinai, 1 Gustave L. Levy Pl, New York, NY 10029, USA; Department of Medicine, Icahn School of Medicine at Mount Sinai, 1 Gustave L. Levy Pl, New York, NY 10029, USA; Digital Health Center at Hasso Plattner Institute, University of Potsdam, Professor-Dr.-Helmert-Str 2–3, 14482 Potsdam, Germany

## Abstract

**Objective:** *Robust phenotyping of patient data from electronic health records (EHRs) at scale is a current challenge in the field of clinical informatics. We introduce Phe2vec, an automated framework for disease phenotyping from EHRs based on unsupervised learning, and we assess its effectiveness against standard rule-based algorithms from the Phenotype KnowledgeBase (PheKB)*.

**Materials and Methods:** *Phe2vec is based on pre-computing embeddings of medical concepts and patients’ longitudinal clinical history. Disease phenotypes are then derived from a seed concept and its neighbors in the embedding space. Patients are similarly linked to a disease if their embedded representation is close to the phenotype. We implemented Phe2vec using 49,234 medical concepts from structured EHRs and clinical notes from 1,908,741 patients in the Mount Sinai Health System. We assessed performance on ten diverse diseases that have a PheKB algorithm*.

**Results:** *Phe2vec phenotypes derived using Word2vec, GloVe, and Fasttext embeddings led to promising performance in disease definition and patient cohort identification with respect to phenotypes and cohorts obtained by PheKB. When comparing Phe2vec and PheKB disease patient cohorts head-to-head using chart review, Phe2vec performed on par or better in nine out of ten diseases in terms of positive predictive values*.

**Discussion:** *Phe2vec offers a solution to improve time-consuming phenotyping pipelines. Differently from other approaches in the literature, it is data-driven and unsupervised, can easily scale to any disease and was validated against widely adopted expert-based standards*.

**Conclusion:** *Phe2vec aims to optimize clinical informatics research by augmenting current frameworks to characterize patients by condition and derive reliable disease cohorts*.

## 1. Objective

Building cohorts for observational experiments requires the reliable identification of patients with the disease of interest. This is difficult to achieve with electronic health records (EHRs) because of data fragmentation and lack of specific inclusion criteria. Diagnoses, in fact, can stem from many forms: documented in a chart note by a physician, in International Classification of Diseases, 9th and 10th revision (ICD-9/10) codes, or as results of a lab test. Depending on the disease, varying data modalities can be better or worse at reflecting reliable diagnosis [1]. For example, a majority of patients with atrial fibrillation are identifiable from their medications, whereas for patients with rheumatoid arthritis, medications are far less useful for classification. Input errors, coding and reporting biases, data availability, sparsity and data structure also present further challenges to accurately identifying patient cohorts [2].

Acknowledging these challenges, EHR-based phenotyping is a computational task to identify key medical concepts in the patient data that consistently and robustly define a disease from EHR data. This is commonly done by applying rule-based algorithms that specify the inclusion or exclusion of certain ICD codes, ranges of laboratory tests, certain medication prescriptions, or the presence of phrases in clinical notes. Phenotyping algorithms are manually built by researchers with advanced knowledge of the specific disease or phenotype of interest, and require validation through manual chart review by experts [3] before being deposited in the Phenotype KnowledgeBase (PheKB) [4]. The Electronic Medical Records and Genomics (eMERGE) consortium led the effort in defining, implementing, and validating such algorithms at various institutions for a number of diseases [5,6]. While effective, implementing a PheKB algorithm on a new dataset is time-intensive as it requires data of varying formats and specific laboratory or clinical information. They also have limited scalability due to the nature of their curated design based on expert knowledge and for a single disease at a time. Consequently, the number of diseases that have public PheKB algorithms is limited, with only 46 diseases or syndromes represented as of July 2020 [7].

Automated phenotyping provides a more rapid and scalable alternative if it can achieve the same robustness as rule-based algorithms [8]. Previous work in this domain used supervised and unsupervised machine learning to derive phenotypes for several diseases, with different strengths and limitations [9–22]. Supervised models rely on classifiers based on manually-labeled gold standards for each specific disease, which is time-consuming and not scalable. Unsupervised approaches discover phenotypes purely from the data, trying to aggregate medical concepts commonly appearing together in the patient records. While more scalable, these approaches are difficult to tune, often rely on defining in advance the number of phenotypes, require manual reviews of the disease definitions and might fail to capture co-occurrences related to less frequent diseases. While innovative and promising, these works were generally not benchmarked against gold standard phenotyping algorithms, i.e., PheKB, to appropriately assess their reliability for identifying cohorts of patients associated with a disease.

This paper presents Phe2vec, a scalable unsupervised learning framework based on neural networks for EHR-based phenotyping. Phe2vec derives vector-based representations, i.e., embeddings, of medical concepts to define the disease phenotypes using the semantic closeness in the embedding space to a seed concept (e.g., an ICD code) [17]. Embeddings are then aggregated at the patient-level to identify populations related to a specific disease based on distance from the phenotype in the embedding space. Experiments based on manual chart review show that Phe2vec performs, at least, as well as PheKB for different and diverse diseases. To the best of our knowledge, this is one of the first head-to-head comparisons between an automated phenotyping method and clinically widely-used rule-based algorithms. Based on unsupervised embeddings of medical concepts, Phe2vec can also potentially be leveraged as the first layer of clinical predictive learning systems and can be extended to include other modalities of data, leading to phenotypes related to a holistic view of the diseases.

## 2. Materials and Methods

This section presents Phe2vec, briefly reviews its underlying methodology, and describes the pipeline implemented for evaluation and comparison.

### 2.1. Phe2vec

**Figure 1** summarizes the conceptual framework of Phe2vec: an automated phenotype algorithm that creates low-dimensional representations (i.e., embeddings) of the medical concepts from longitudinal EHRs [17]. These representations put all concepts from both structured and unstructured EHRs in a common phenotype space where association is inversely proportional to pairwise distance (**Fig. 1A**). A disease phenotype is defined as a seed concept and its neighborhood. Embeddings are then used to summarize patient history and measure their relatedness with the phenotype using distance analysis (**Fig. 1B**).

**Figure 1:**
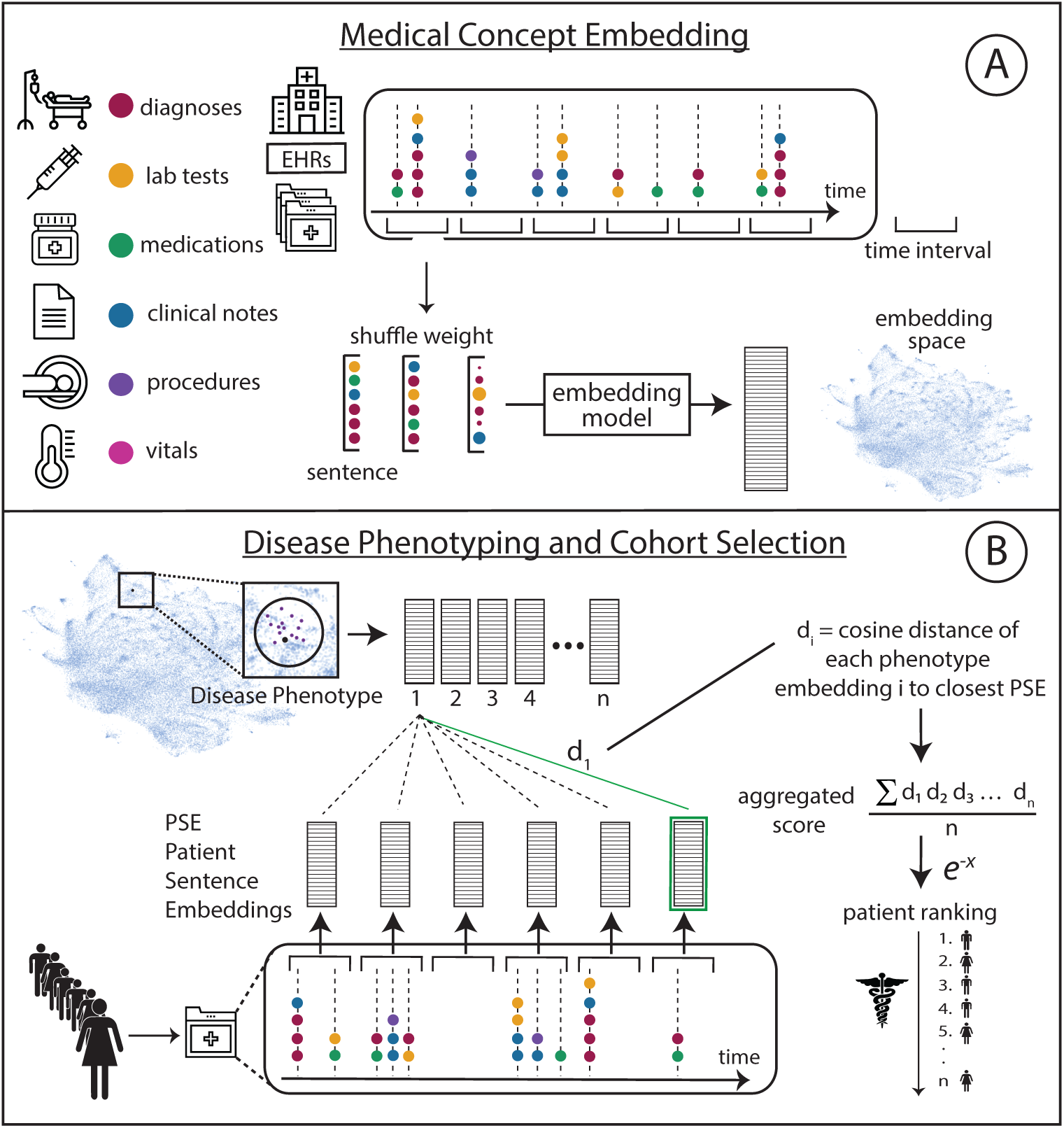
Phe2vec framework. (A) An embedding algorithm creates low-dimensional vector-based representations of medical concepts from longitudinal EHRs. (B) Disease phenotypes are defined by considering a seed concept (e.g., an ICD code) and its neighbors in the embedding space. A patient’s clinical history is summarized by aggregating all the medical concept embeddings. This representation is used to measure the distance of the patient with the phenotype in the vector space to determine the association with the disease.

#### Medical Concept Embeddings

Longitudinal EHRs are irregularly-sampled temporal sequences of medical concepts. Concepts adjacent to each other in these sequences should group together in the learned embedding space. To this aim, we first partition the patient data time intervals composed by *N* days. Second, we remove duplicates from each time interval and third, we randomly shuffle the concepts in each interval [23]. This process is done to reduce biases related to how the data are inserted in the system. Each time interval represented as a sequence of unique medical concepts is then considered as a “sentence” (where each medical concept is a “word”) and can be modeled, e.g., using embedding algorithms from the NLP literature, such as Word2vec [24], GloVe [25] and FastText [26]. Regardless of the specific algorithm, after training, every medical concept is represented as a low-dimensional vector, with all the medical concepts mapped in the same metric space.

#### Definition of Disease Phenotypes

The disease phenotype is defined from medical concept embeddings by exploring the neighborhood of a specific seed query, for example the ICD code that is related to the specific condition. The size of the neighborhood can be tuned differently based on the disease but, in order to reduce noise, is limited to the concepts within a certain distance from the seed concept.

#### Patient Representation

Patient clinical histories are summarized by aggregating medical concept embeddings. In particular, for each time interval in the patient clinical history (i.e., the “sentence”), we compute the weighted average of all medical concept embeddings within that sentence and subtract the projections of the average vectors on their first principal component [27]. This serves to remove the largely shared components from the vectors, leading to more discriminative aggregated representations. The weight of each medical concept *w* is defined as *1 / (1 + p(w))*, with *p(w)* being the estimated medical concept frequency across the dataset, leading to lower weights for frequent medical concepts. Every patient is then characterized by a sequence of aggregated embeddings, one per each time interval, lying in the same space of the medical concepts. We refer to these aggregated embeddings as “patient sequence embeddings” (PSE).

#### Automated Definition of Disease Patient Cohorts

To quantify the association between a patient and a disease, we compute the distance between each medical concept in the disease phenotype and each PSE in the patients’ clinical history. Then, for each concept in the phenotype, we take the closest PSE and average all these distances to obtain the aggregated score. Lastly, the latter is transformed to an adjacency measure by applying the inverse of the exponential function. We refer to this aggregated score as “phenotype score”.

### 2.2. PheKB: a Phenotype KnowledgeBase

PheKB is a collection of phenotyping algorithms built by experts for a variety of diseases, which are widely considered the clinical gold standards of EHR-based phenotyping. This effort was initiated within the eMERGE Network in 2012 and is continuously enhanced as needs are identified and the field advances [4]. Each algorithm is composed by a manually built set of inclusion and exclusion criteria that are used to define a certain condition and to identify patients within a cohort of EHRs. The data types commonly included in the rules are related to ICD codes, medications, Current Procedural Terminology, version 4 (CPT-4) procedure codes, descriptions for laboratory tests and vital signs, demographic information such as age and race, and words and phrases extracted from clinical notes.

After assessing the feasibility of creating a specific disease algorithm within the eMERGE Network, the clinical site develops, implements, and tests the algorithm before sharing it on PheKB for iterative feedback and revision. Uploaded documents typically include full descriptions of the computable algorithms including data types used, execution logic and variable dependencies, data definitions, performance metrics, and descriptive graphics. Each algorithm’s performance is validated within different clinical repositories for establishing transportability to a large number of collaborating sites. Once the algorithm is marked as “final” by the author, it is made available to the whole community. Researchers in other institutions can then follow the logic and rules defined to retrieve cohorts of patients in their EHRs.

### 2.3. Evaluation Design

**Figure 2** highlights the study design to compare Phe2vec and PheKB on different diseases using EHRs from a large-scale hospital system.

**Figure 2:**
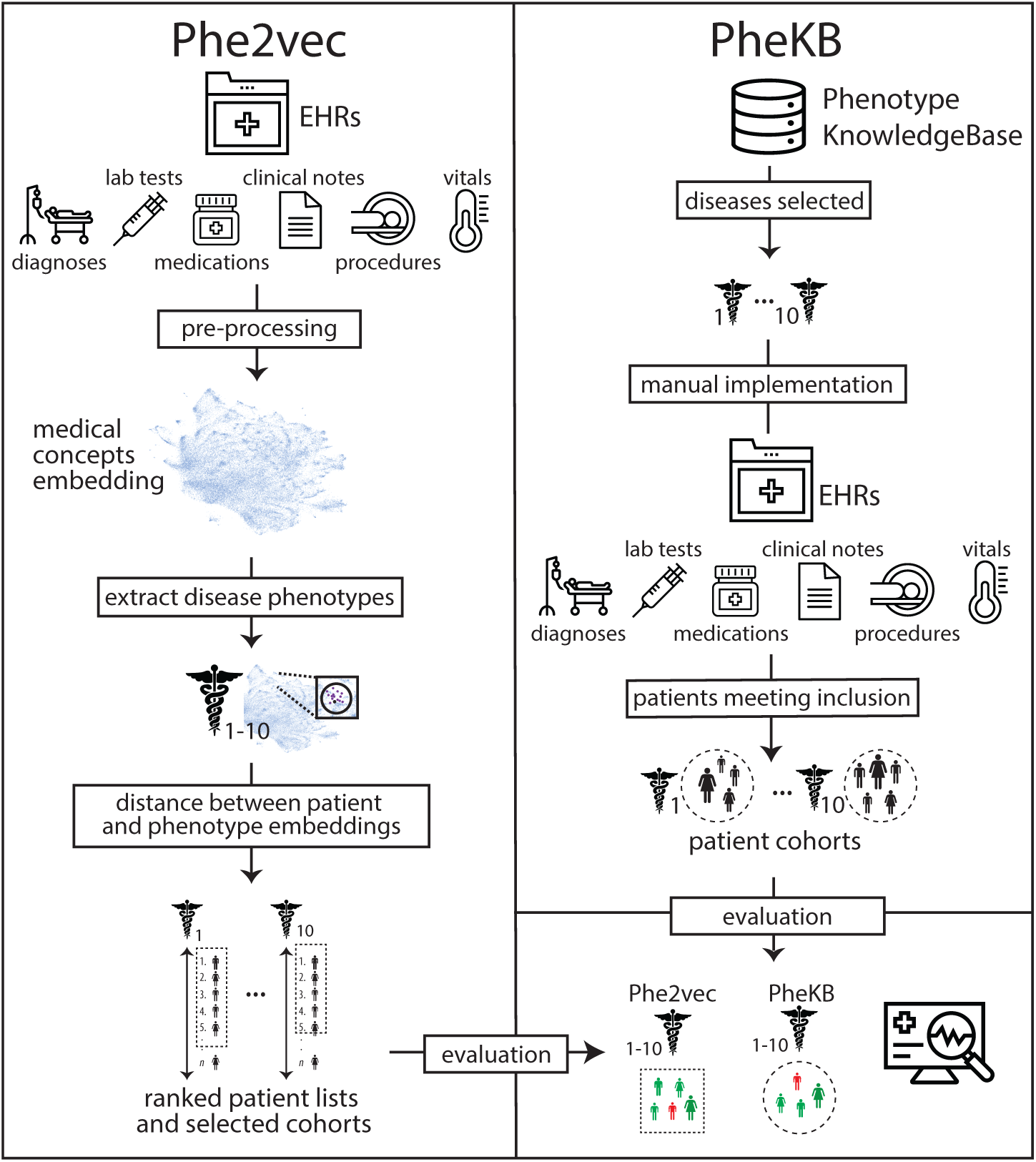
Study design to compare Phe2vec, an automated phenotyping algorithm, to PheKB, a bank of manually-derived phenotyping rules, on different diseases using EHRs from a large-scale hospital system.

#### Dataset

We used de-identified EHRs from the Mount Sinai Health System (MSHS) data warehouse; the study was approved under IRB-19-02369 by the Program for the Protection of Human Subjects at the Icahn School of Medicine at Mount Sinai. MSHS is a large and diverse urban hospital located in New York, NY, which generates a high volume of structured, semi-structured, and unstructured data from inpatient, outpatient, and emergency room visits. Patients in the system can have up to 15 years of follow-up data unless they are transferred or move their residence away from the hospital system. We accessed a de-identified version of the data containing ∼4.5 million patients, spanning the years from 1980 to 2016.

For each patient, we aggregated ICD-9 diagnosis codes, medications normalized to RxNorm, CPT-4 procedure codes, vital signs, and lab tests normalized to Logical Observation Identifiers Names and Codes (LOINC). ICD-10 codes were mapped back to the corresponding ICD-9 versions. We preprocessed clinical notes using a tool based on the Open Biomedical Annotator to extract clinical concepts from the free text [28,29]. The vocabulary was composed of 57,464 clinical concepts.

We retained all patients with at least two concepts, resulting in a collection of 1,908,741 different patients, with an average of 88.7 concepts per patient. In particular, the cohort included 1,068,940 females, 820,239 males, and 19,562 not declared; the mean age of the population as of 2016 was 48.33 years (s.d. = 23.71). We used 300,000 random patients for training the medical concept embeddings and the remaining 1,608,741 patients for testing. We decided on this split because we wanted to evaluate the phenotype algorithms on retrieving cohorts of patients from a large population.

#### Diseases

We selected diseases from PheKB by filtering publicly available algorithms by “Type of Phenotype” equal to “Disease or Syndrome”. From this set we identified ten heterogenous diseases for the evaluation: abdominal aortic aneurysm (AAA), atrial fibrillation, attention deficit hyperactivity disorder (ADHD), autism, Crohn’s disease, dementia, herpes zoster, multiple sclerosis, sickle cell disease, and type 2 diabetes mellitus (T2D). We selected diseases with algorithms that could be implemented with the MSHS data available for this project and were well represented in the dataset.

#### Implementation Details

We learned medical concept embeddings using the 300,000 patients in the training set. We tested a large number of configurations (e.g., time interval N ranging from 3 to 60 days; embedding dimensions spanning from 10 to 1,000; minimum concept frequency from 2-10). We trained different embeddings using Word2vec, GloVe, and FastText. We trained Word2vec and FastText with skip-gram and negative sampling [30], while GloVe was trained with the standard configuration [25]. We optimized hyperparameters of all models by measuring the clinical relevance of the neighbors in the embedding space of all ICD codes in the vocabulary [23]. In particular, we used the Clinical Classification System (CCS), single level, to group ICD codes into higher-level clinically meaningful categories. We then evaluated whether and to which extent the nearest neighbors of each ICD code included other ICD codes from the same CCS group. For brevity here we report only the best setting derived from this hyperparameter optimization which was then used in the rest of the evaluation.

For each patient trajectory, we used time intervals of N=15 days and retained all concepts appearing at least three times. We obtained embeddings with size equal to 200 for 49,234 medical concepts. Each patient in the test set was then summarized as a sequence of PSEs covering non-empty 15 days intervals along the clinical trajectory. We used cosine distance to measure relationships in the phenotype space for both medical concepts and patients.

PheKB algorithms were implemented for the ten diseases selected by including in the phenotypes all the medical concepts specified for each disease and that were available in the dataset (**Supplementary Table 1**). We were able to successfully implement all algorithms with only a few minor specifications. More details about the implementations are available in the Supplementary Material. While some of these algorithms include inclusion criteria for a control group, we focused only on case selection. For algorithms that differentiated cases into several types, we simply aggregated all types. The medical concepts associated with each disease phenotype are reported in **Supplementary Table 2**.

#### Phenotype Evaluation

For each disease we defined Phe2vec phenotypes by starting from the associated ICD code (**Supplementary Table 3**) and retaining the top K closest concepts in the embedding space (i.e., “neighbors”). We then ranked patients based on their distance with such phenotype definitions. PheKB patients were retrieve by simply considering all the patients satisfying the logic defined in each disease phenotype. These algorithms do not specify a score, consequently patients were not ranked and were treated as equally associated with the disease.

## 3. Results

We compared Phe2vec and PheKB in terms of phenotype definition and automated disease cohort selection.

### Disease Phenotype Analysis

To assess the performance of Phe2vec in building phenotypes, for each disease, we evaluated the overlap between the medical concepts retrieved from the seed neighbors and those in PheKB. Since the number of concepts in a PheKB definition (i.e., recall level R) varies across diseases, we evaluated different neighbors with sizes equal to percentages of R of the corresponding disease (R%) [18]. We measured precision and recall per disease, where precision is the number of correct positive results divided by the number of all positive results, and recall is the number of correct positive results divided by the number of positive results that should have been returned. Results averaged over the ten diseases are reported in **Figure 3**. Overall, phenotypes obtained with Word2vec as an embedding algorithm led to better performances compared to GloVe and FastText. At recall level (R% = 100), Word2vec obtained an F-score (harmonic mean of precision and recall) equal to 0.41, compared to 0.28 and 0.38 of GloVe and FastText, respectively. The phenotypes most related to PheKB were obtained for autism, dementia and sickle cell disease, while AAA and herpes zoster obtained the worst results.

**Figure 3:**
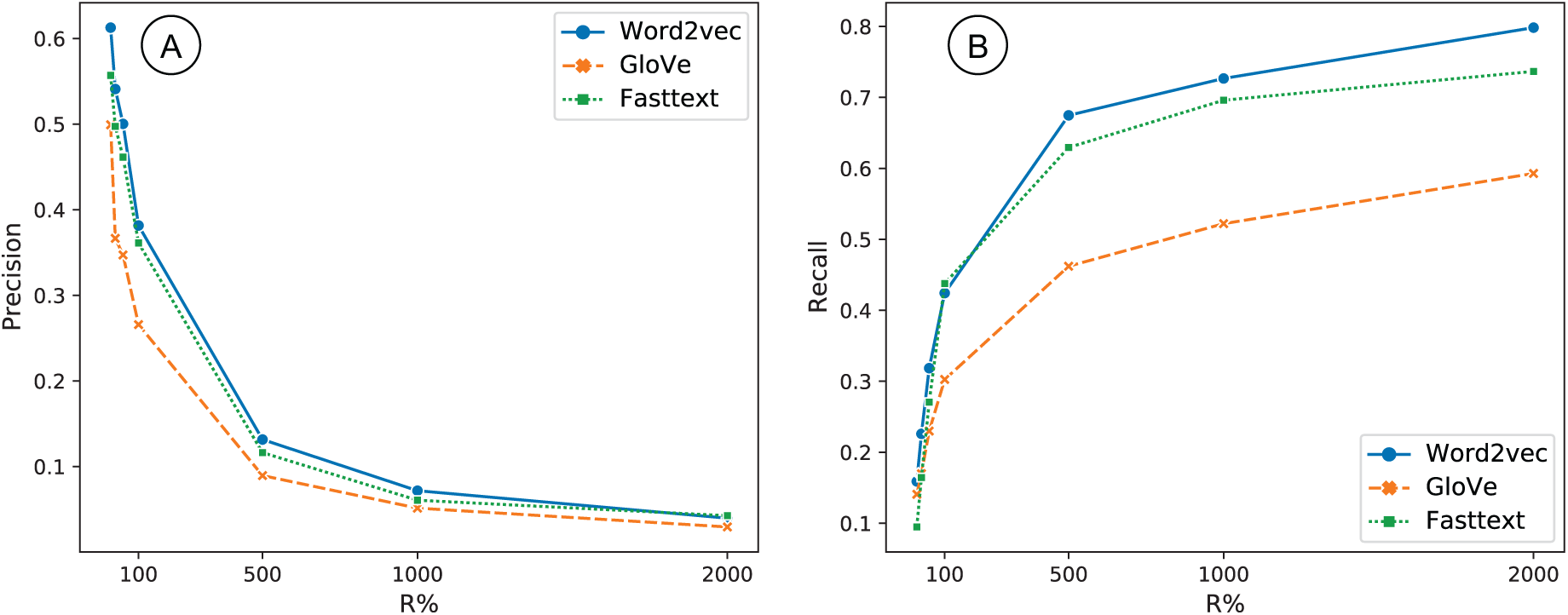
Precision (A) and recall (B) obtained by phenotypes derived with Phe2vec when matched against PheKB averaged over the diseases. For each disease, we considered different neighborhoods of the corresponding seed concept with sizes equal to percentages of the recall level (R%), which is the number of concepts in the PheKB phenotypes.

**Figure 4** visualizes the (a) phenotype space generated by Phe2vec with Word2vec embeddings using UMAP (Uniform Manifold Approximation and Projection for dimension reduction [31]) and highlights the phenotypes created for (b) T2D and (c) dementia. See **Supplementary Figure 1** for the phenotype space of the other diseases considered. As it can be seen, most of the concepts are clinically related to the condition and create a “disease definition” that can be used to better identify cohorts of case/control patients in the dataset.

**Figure 4:**
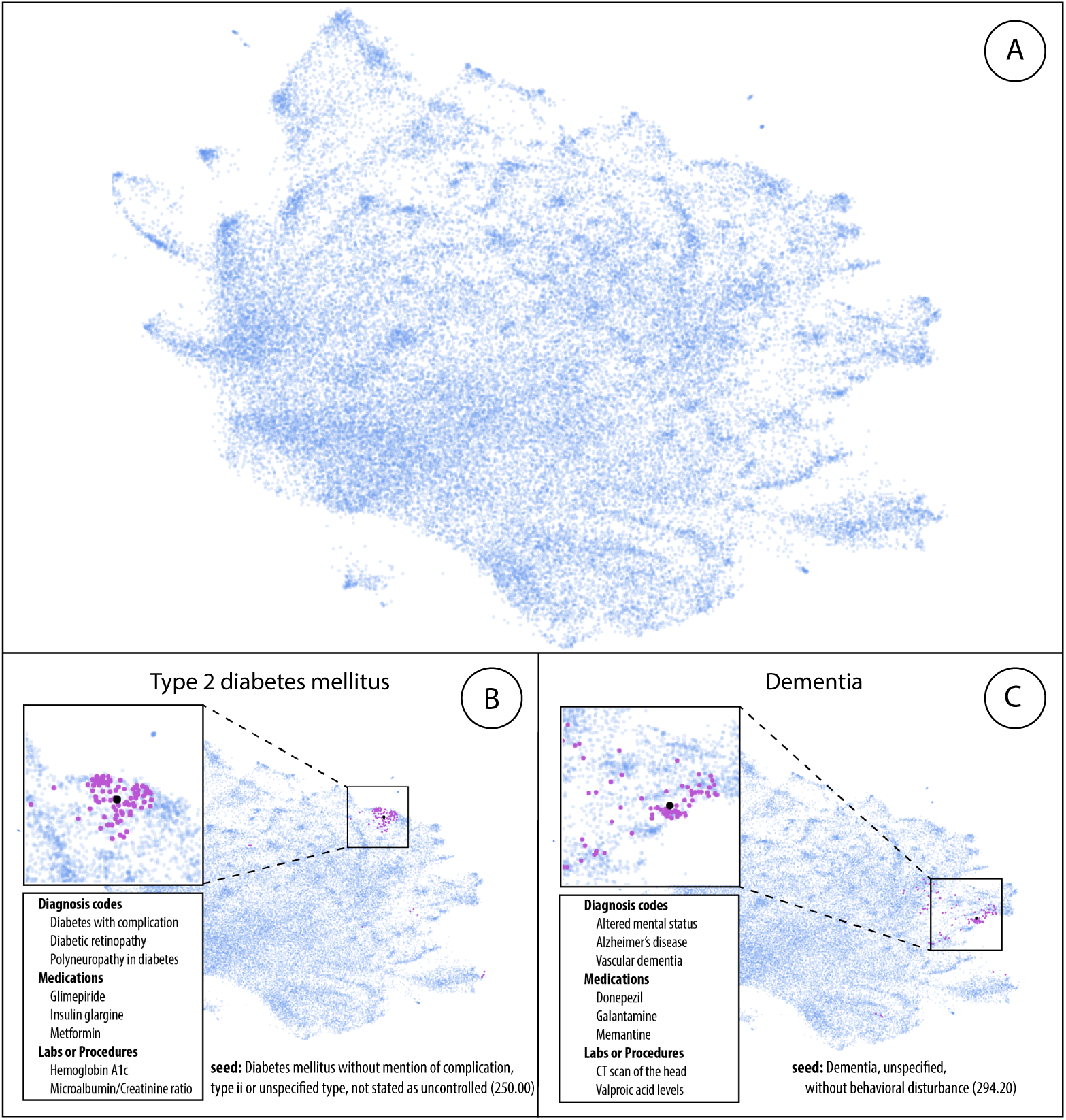
Uniform Manifold Approximation and Projection (UMAP) visualization of the EHR-based phenotype space generated by Phe2vec with Word2vec embeddings (A). Phenotypes for type 2 diabetes mellitus (B) and dementia (C). Seed concepts are colored in black, while concepts in the phenotypes are colored in purple.

### Disease Cohort Selection

We used phenotypes based on different embeddings to retrieve cohorts of patients for each disease. We defined a disease phenotype by retaining the seed concept and its closest neighbors. In a practical scenario, a domain expert would choose the optimal number of concepts. Here, for the sake of generalization, we retained concepts with phenotype scores greater than 0.7. This value was chosen to reduce noise in the phenotype definition, while still including at least five concepts per disease. We computed phenotype scores between patients and diseases, and we evaluated annotation and retrieval performances against cohorts retrieved with PheKB. We included in the experiment an approach based on a bag of concepts (“BoCon”), which, for each patient, simply counts the occurrence of each concept in the phenotype identified by Phe2vec (rather than measuring diseases in the embeddings space). We also assess the performance of two commonly used phenotyping methods, PheCode [22] and PheMap [21]. PheCode groups ICD-9/10 codes into clinically meaningful phenotypes, thereby collapsing the diagnosis code space. PheMap is a high-throughput phenotyping approach that identified concepts important to phenotypes from publicly available sources like MedlinePlus, MedicineNet, and Wikipedia. We implemented both of these methods to retrieve cohort of patients for each disease and likewise evaluated annotation and retrieval performance against cohorts retrieved with PheKB. Results averaged over the ten diseases are presented in **Table 1**. The annotation experiment relies on a threshold to discriminate between “phenotype” vs. “non-phenotype”, with scores greater than this threshold identifying patients with the phenotype of interest. In order to choose a value independently and ensure generalizability, for this task we organized a 2-fold cross-validation experiment where we randomly split the dataset in half, obtaining two independent cohorts of ∼800,000 patients that we used to train and test the threshold (and vice versa). During training, for each disease, we ranged the value from 0.1 to 1, with 0.05 increments, and retained the threshold leading to the best results across all diseases in the training set. We then applied that value to the corresponding test sets to annotate patients with the phenotypes and evaluate results in terms of F-score averaged across the two folds. For BoCon, PheCode and PheMap we annotated the disease for all patients with at least one concept from the corresponding phenotype. In the retrieval experiment, for each disease, we sorted all 1.6M patients by the distance with the phenotype and measured the position in the ranking of the PheKB patients. We report precision at the recall level (R-precision) and the area under the precision and recall curve (AUC-PR). R-precision measures the number of positive patients in the top R position of the rank, where R is the number of true patients associated with the disease. The PR curve is a plot of precision and recall for different thresholds; AUC-PR is computed by integrating the PR curve. As seen, methods based on Phe2vec outperforms BoCon as well as PheCode and PheMap for all metrics and embeddings. Using phenotypes composed by multiple medical concepts leads to better results than simply using the seed concept. While expected, this indicates the need of inclusive phenotypes methods that overcome limitations of ICD codes (as also indicated by PheCode performances). As in the previous experiment, Phe2vec based on Word2vec overall obtains slightly better results than using other embedding models. This setting also obtains statistically significant better results than BoCon (t-test, p=0.05). From now on we will generally refer to this configuration as “Phe2vec”. **Table 2** shows results obtained for each disease using Phe2vec (see **Supplementary Tables 4-6** for the results obtained using GloVe, FastText and BoCon, respectively).

**Table 1:** Disease cohort selection results obtained with the automated evaluation, where PheKB cohorts are considered as gold standard. Cohorts are retrieved using a unique seed ICD code, or the corresponding disease phenotype obtained with Phe2vec. We compare embedding-based methods (Word2vec, GloVe, FastText), which rely on distance between patients and phenotypes, bag of codes (BoCon), which just count the frequency of the phenotype concepts in the patient history PheCode [21], and PheMap [21]. All results are average across ten diseases.

|  |  | **F-score** | **R-precision** | **AUC-PR** |
| --- | --- | --- | --- | --- |
|  | PheCode | 0.44 | 0.41 | 0.53 |
|  | PheMap | 0.49 | 0.42 | 0.55 |
| *Seed Code* | BoCon | 0.42 | 0.39 | 0.53 |
|  | Word2vec | 0.50 | 0.52 | 0.59 |
|  | GloVe | 0.43 | 0.51 | 0.54 |
|  | FastText | 0.47 | 0.50 | 0.58 |
| *Disease*<br>*Phenotype* | BoCon | 0.53 | 0.44 | 0.56 |
|  | Word2vec | **0.64** | **0.62** | **0.69** |
|  | GloVe | 0.57 | 0.54 | 0.62 |
|  | FastText | 0.59 | 0.55 | 0.64 |

**Table 2:** Results on cohort selection per disease obtained by Phe2vec, with Word2vec embeddings, where PheKB cohorts are considered as gold standard.

| Disease | Patients | F-score | R-precision | AUC-PR |
| --- | --- | --- | --- | --- |
| Abdominal aortic aneurysm | 1,982 | 0.64 | 0.57 | 0.73 |
| Attention deficit hyperactivity disorder | 7,778 | 0.72 | 0.62 | 0.77 |
| Atrial fibrillation | 39,568 | 0.54 | 0.54 | 0.56 |
| Autism | 1,279 | 0.53 | 0.57 | 0.58 |
| Crohn's disease | 6,207 | 0.73 | 0.69 | 0.78 |
| Dementia | 15,406 | 0.58 | 0.58 | 0.58 |
| Herpes zoster | 1,618 | 0.45 | 0.46 | 0.57 |
| Multiple sclerosis | 4,532 | 0.85 | 0.82 | 0.86 |
| Sickle cell disease | 949 | 0.69 | 0.75 | 0.71 |
| Type 2 diabetes mellitus | 59,233 | 0.65 | 0.59 | 0.73 |

We lastly compared Phe2vec and PheKB head-to-head using manual review to assess their performances independently. For each of the ten disease cohorts, we selected 50 PheKB-identified patients and 50 Phe2vec-identified patients, resulting in 100 patients per disease. We performed manual chart review to identify whether the targeted disease diagnosis was explicitly given at any time in any clinical note for each given patient. This process consisted of randomly assigning each of these 1,000 cases to two of three possible raters who would then read over each individual’s notes from all encounters in order to find a diagnosis for the particular disease. Each rater evaluated their assigned individuals independently and was blinded to algorithm predictions. If the two raters agreed on whether the patient had the disease or not, that would be the true disease status label. In cases of disagreement, a decision was reached by a second round of review with a consensus reached by all raters, so that each patient’s disease status was confirmed by at least two, and up to three raters. **Supplementary Figure 2** shows the chart review inter-rater reliability following the point at which individual raters had made their first assessments.

**Table 3** reports results for all ten diseases in terms of positive predictive value (PPV), which is the proportion of patients marked as positive by the algorithm that truly has the disease as defined by chart review. Phe2vec obtained better PPV in nine diseases, with highest improvements for herpes zoster and T2D, showing qualitative performances on-par with manual phenotypes. Overall, Phe2vec and PheKB achieved an average PPV of 0.94 and 0.82, respectively.

**Table 3:** Per-disease positive predictive value (PPV) obtained by Phe2vec and PheKB against a gold standard derived via manual chart review of progress notes.

| Disease | Positive Predictive Value |  |
| --- | --- | --- |
|  | Phe2vec | PheKB |
| Abdominal aortic aneurysm | 1.00 | 0.95 |
| Attention deficit hyperactivity disorder | 0.97 | 0.85 |
| Atrial fibrillation | 0.85 | 0.85 |
| Autism | 0.81 | 0.95 |
| Crohn’s disease | 0.98 | 0.81 |
| Dementia | 0.98 | 0.82 |
| Herpes zoster | 0.92 | 0.45 |
| Multiple sclerosis | 0.97 | 0.97 |
| Sickle cell disease | 0.96 | 0.83 |
| Type 2 diabetes mellitus | 0.98 | 0.74 |

## 4. Discussion

This study proposes a computational framework based on unsupervised machine learning to define disease phenotypes from heterogeneous EHRs. Specifically, we developed and validated an architecture named Phe2vec that infers informative vector-based representations of medical concepts and uses distance analysis from a seed concept to define phenotypes and to retrieve cohorts of patients associated with diseases. Phe2vec aims to be domain-free, robust and scalable to all diseases. Experimental results on large-scale EHRs show that Phe2vec identifies similar phenotypes to PheKB and can be used to accurately identify cohorts of patients diagnosed with a certain disease. In particular, Phe2vec performed on par or outperformed PheKB algorithms in nine out of ten diseases examined. Experiments also highlight the slight preference of Word2vec as a model to learn medical concepts embeddings from EHRs over Fasttext and Glove. Phe2vec is purely data driven and requires no manual effort beyond the selection of a single seed concept, which can simply be the general ICD code associated with the targeted disease. We release the 100 most related medical concepts from Phe2vec for each ICD-9 code at: https://github.com/HPIMS/phe2vec.

### Background and Significance

Research based on EHRs is central to fulfilling the vision of personalized medicine [32– 40]. However, due to their nature of being structured for billing purposes, carefully designing scientific experiments is difficult. Advanced EHR phenotyping of diseases is best understood as an “expert system”, where researchers with advanced knowledge of a particular disease design a list of criteria which may be used to identify affected (i.e., cases) and sometimes assuredly non-affected (i.e., controls) individuals typically by intricate rule-based algorithms specifying the presence and/or absence of particular billing codes, pre-defined ranges for laboratory tests, prescription of characteristic medications, processing of clinical notes, among others [4]. However, these approaches are time-consuming, and several machine learning phenotyping methods have been proposed to speed-up automatic phenotype definition. Supervised approaches that require domain experts to label datasets is a clear bottleneck, especially in medicine, due to time and difficulty of collecting reliable annotations [16]. In the “learning with anchors” framework, the authors propose a semi-supervised method where observations with high positive predictive value for the phenotypes of interest are identified by domain experts, and these are then used to learn labels for the dataset and to train classifiers to identify phenotypes [13]. While more scalable, this method still relies on manual experts to define observations from the raw data. Phe2vec, differently, is an unsupervised framework where human supervision is minimum and required, in case, only at the end to optimize the automated phenotype definitions (thus limited to a small subset of concepts). Unsupervised learning methods such as latent Dirichlet allocation [12,19] and tensor factorization [10,11,15] learn phenotypes without the need of human supervision [21,22]i. These methods showed promising results but rely on modeling bags of medical concepts, without considering the inherent temporality of patient EHRs. Additionally, they require specification of the number of phenotypes in advance (as they aim to discover pre-fixed latent groups of medical concepts frequently co-occurring in the data), so they are not very flexible, and need human evaluation to assess clinical relevance of all phenotypes (i.e., define what each phenotype is referring to). This is time-consuming and makes it difficult to scale to large datasets covering many diverse diseases. Alternatively, Phe2vec uses pre-trained embeddings to derive phenotypes and can scale to all diseases by requiring the definition of one seed concept. This leads to a flexible solution that can evolve through time and can be extended and applied to other institutes as well. Lee et al. explored the use of embeddings based on GloVe and graph-modeling as well obtaining similar results in terms of phenotype overlap with PheKB [41]. While they included more diseases (33) in the experiments, they did not evaluate the phenotypes in retrieving patients, which is the primary motivation for disease phenotyping. Moreover, they modeled global co-occurrences of medical concepts across the data, without considering the longitudinality of clinical data. Phe2vec extends our previous work [17] by including clinical notes, a larger cohort of patients and diseases, and including manual chart review comparing Phe2vec and PheKB and the feasibility of using Phe2vec to create phenotypes for diseases not covered in PheKB. To the best of our knowledge, this is the first work providing a thorough head-to-head comparison between an automated phenotype algorithm and PheKB.

### Potential Applications

Phe2vec provides a framework to automatically derive disease phenotypes from EHRs. Based on medical concept embeddings derived from a large-scale heterogeneous EHR dataset, this approach promises to be easily deployable in other facilities with minimum effort. Additionally, these embeddings can be used to initialize machine learning architectures for clinical predictive analysis and medical research [40,42].

The natural application is to identify medical concepts related to a disease diagnosis and use them to identify reliable cohorts of patients for case-control studies. An automated method such as Phe2vec can be used as a stand-alone tool in clinical facilities but can also be used to improve and scale the creation of PheKB definitions. In fact, domain-experts can use Phe2vec to quickly generate a list of candidate medical concepts, manually refine them, evaluate them in a multi-center scenario and release it as standards. This would considerably speed up operations and would provide a larger number of phenotypes available as standards. Data-driven phenotypes derived automatically and updated constantly from EHRs can also help identify changes in clinical practice and guidelines. This could ultimately increase or decrease the significance of some concepts as well as introduce new diagnostic lab tests or medications.

Phe2vec aims to contribute to the next generation of clinical systems that can scale to millions of patient records and use machine learning to effectively support clinicians in their daily activities. The ability to quickly derive disease phenotypes in the EHRs for a large number of diseases can be used to easily track clinical history of patients. EHRs are typically organized by data type, with medical problems, medications, procedures, and lab results chronologically sorted in separate areas of the chart. As a result, it can be difficult to aggregate all the relevant information to define previous conditions and current clinical status [43]. Large-scale disease phenotypes can provide an alternative to organize medical records by condition with all pertinent information grouped together. In this scenario, individuals’ clinical histories are summarized by previous and current diseases, facilitating the contextualization of patients when they return to the hospital.

### Limitations

The main goal of this work was to prove feasibility and robustness of Phe2vec in comparison to PheKB. There are several limitations to our study. First, we acknowledge the use of laboratory test presence only and not of the test result values themselves. While test frequency is often sufficient when modeling large data sets of patients and for a number of diseases [44], result values might be necessary for some other diseases and should be included. To this point, for example, lab results can be categorized into discrete values (e.g., “high”, “normal”, “low”) and aggregated into extended concepts combining test and associated result (e.g., “<lab_test>|<lab_result_category>“). Second, the extensive amount of time to implement PheKB algorithms and for chart review prevented us from including more disease categories in the experiment. A larger scale evaluation, including validation within other hospital systems, is required before deployment in any clinical practice. In the cohort retrieval experiments, we defined the phenotypes using an arbitrary closeness threshold. In practice, a domain-expert should manually revise the list of medical concepts in the phenotype and choose the appropriate cut-off level. Definition of an automated method to select the optimal neighborhood of the seed concept would increase scalability and reduce this human intervention. Additionally, due to the time intensive nature of the manual chart review process, we were limited from performing in depth error analysis, which could elucidate common reasons for mistakes. Alternative strategies for arriving at these labels could identify medical concepts associated with misclassification. Lastly, we only considered the use of ICD-9 codes as seed concepts. Using more specific diagnosis, medications, lab tests, or a combination of codes might change phenotype definitions and improve performances.

### Future Work

Future work will attempt to address these limitations and to further enhance and replicate the framework. To start, we plan to evaluate more sophisticated methods to summarize patient trajectories. While a weighted average of medical concepts was enough to show effectiveness of Phe2vec, architectures based on unsupervised deep learning better represent patient clinical histories and promise to improve modeling the interactions between patients and phenotypes [40,45,46]. Next, we will evaluate other strategies to create medical concept embeddings, such as the use of transformer architectures to model both clinical notes and structured EHRs [47,48]. Third we will define a framework to analyze how phenotypes change over time with the goal of improving disease definitions and their association with patients. We will also explore the use of Phe2vec to create reliable disease-specific control cohorts for observational studies. Finally, we will embed other modalities of data, such as genetics and clinical imaging, into this framework, which should refine disease phenotypes and potentially reveal novel associations.

## 5. Conclusion

We introduced Phe2vec, an automated method based on unsupervised machine learning for EHR-based disease phenotyping. Phe2vec uses embeddings of medical concepts to derive phenotypes and to measure the association between disease and patient representations. We obtained results that are comparable with electronic phenotyping algorithms that use manually defined rules from PheKB. Automated architectures for disease phenotype that are capable of scaling to a large number of diseases, patients, and health data promise to offer a more holistic way to examine disease complexity and to improve clinical practice and medical research.

## Data Availability

n/a

## Contributions

R.M. and B.S.G. initiated the idea. R.M. collected the data, conducted the research and the experimental evaluation and wrote the manuscript. J.K.D.F., K.W.J., and B.S.G. implemented the PheKB algorithms, performed the manual chart review of the results and refined the article; G.N.N. provided clinical support and refined the article; E.G. refined the article; J.T.D. and E.P.B. supported the research. All the authors edited and reviewed the manuscript.

## Code Availability

The code is available at: https://github.com/HPIMS/phe2vec.

## References

1 Wei W-Q, Teixeira PL, Mo H, et al. Combining billing codes, clinical notes, and medications from electronic health records provides superior phenotyping performance. J Am Med Inform Assoc 2016;23:e20–7.

2 Weiskopf NG, Weng C. Methods and dimensions of electronic health record data quality assessment: enabling reuse for clinical research. J Am Med Inform Assoc 2013;20:144–51.

3 Pathak J, Kho AN, Denny JC. Electronic health records-driven phenotyping: challenges, recent advances, and perspectives. J Am Med Inform Assoc 2013;20:e206–11.

4 Kirby JC, Speltz P, Rasmussen LV, et al. PheKB: a catalog and workflow for creating electronic phenotype algorithms for transportability. Journal of the American Medical Informatics Association. 2016;23:1046–52. doi:10.1093/jamia/ocv202

5 Gottesman O, and The eMERGE Network, Kuivaniemi H, et al. The Electronic Medical Records and Genomics (eMERGE) Network: past, present, and future. Genetics in Medicine. 2013;15:761–71. doi:10.1038/gim.2013.72

6 Newton KM, Peissig PL, Kho AN, et al. Validation of electronic medical record-based phenotyping algorithms: results and lessons learned from the eMERGE network. J Am Med Inform Assoc 2013;20:e147–54.

7 Public Phenotypes | PheKB. https://phekb.org/phenotypes (accessed 22 Jul 2020).

8 Banda JM, Seneviratne M, Hernandez-Boussard T, et al. Advances in Electronic Phenotyping: From Rule-Based Definitions to Machine Learning Models. Annu Rev Biomed Data Sci 2018;1:53–68.

9 Carroll RJ, Eyler AE, Denny JC. Naïve electronic health record phenotype identification for rheumatoid arthritis. In: AMIA annual symposium proceedings. American Medical Informatics Association 2011. 189.

10 Ho JC, Ghosh J, Steinhubl SR, et al. Limestone: high-throughput candidate phenotype generation via tensor factorization. J Biomed Inform 2014;52:199–211.

11 Wang Y, Chen R, Ghosh J, et al. Rubik: Knowledge Guided Tensor Factorization and Completion for Health Data Analytics. KDD 2015;2015:1265–74.

12 Pivovarov R, Perotte AJ, Grave E, et al. Learning probabilistic phenotypes from heterogeneous EHR data. J Biomed Inform 2015;58:156–65.

13 Halpern Y, Horng S, Choi Y, et al. Electronic medical record phenotyping using the anchor and learn framework. J Am Med Inform Assoc 2016;23:731–40.

14 Chiu P-H, Hripcsak G. EHR-based phenotyping: Bulk learning and evaluation. Journal of Biomedical Informatics. 2017;70:35–51. doi:10.1016/j.jbi.2017.04.009

15 Henderson J, Ho JC, Kho AN, et al. Granite: Diversified, Sparse Tensor Factorization for Electronic Health Record-Based Phenotyping. In: 2017 IEEE International Conference on Healthcare Informatics (ICHI). 2017. 214–23.

16 Yu S, Ma Y, Gronsbell J, et al. Enabling phenotypic big data with PheNorm. J Am Med Inform Assoc 2018;25:54–60.

17 Glicksberg BS, Miotto R, Johnson KW, et al. Automated disease cohort selection using word embeddings from Electronic Health Records. Pac Symp Biocomput 2018;23:145–56.

18 Lee J, Liu C, Kim JH, et al. Comparative Effectiveness of Knowledge Graphs-and EHR Data-Based Medical Concept Embedding for Phenotyping. medRxiv Published Online First: 2020.https://www.medrxiv.org/content/10.1101/2020.07.14.20151274v1.abstract

19 Ahuja Y, Zhou D, He Z, et al. sureLDA: A Multi-Disease Automated Phenotyping Method for the Electronic Health Record. bioRxiv Published Online First: 2020.https://www.biorxiv.org/content/10.1101/2020.04.13.038968v1.abstract

20 Wagholikar KB, Estiri H, Murphy M, et al. Polar labeling: silver standard algorithm for training disease classifiers. Bioinformatics 2020;36:3200–6.

21 Zheng NS, Feng Q, Kerchberger VE, et al. PheMap: a multi-resource knowledge base for high-throughput phenotyping within electronic health records. J Am Med Inform Assoc 2020;27:1675–87.

22 Wu P, Gifford A, Meng X, et al. Mapping ICD-10 and ICD-10-CM codes to phecodes: Workflow development and initial evaluation. JMIR Med Inform 2019;7:e14325.

23 Choi Y, Chiu CY-I, Sontag D. Learning Low-Dimensional Representations of Medical Concepts. AMIA Jt Summits Transl Sci Proc 2016;2016:41–50.

24 Mikolov T, Chen K, Corrado G, et al. Efficient Estimation of Word Representations in Vector Space. arXiv [cs.CL]. 2013.http://arxiv.org/abs/1301.3781

25 Pennington J, Socher R, Manning CD. Glove: Global vectors for word representation. In: Proceedings of the 2014 conference on empirical methods in natural language processing (EMNLP). 2014. 1532–43.

26 Bojanowski P, Grave E, Joulin A, et al. Enriching Word Vectors with Subword Information. Transactions of the Association for Computational Linguistics 2017;5:135–46.

27 Arora S, Liang Y, Ma T. A Simple but Tough-to-Beat Baseline for Sentence Embeddings. 2016.https://openreview.net/pdf?id=SyK00v5xx (accessed 22 Jul 2020).

28 Jonquet C, Shah NH, Musen MA. The open biomedical annotator. Summit Transl Bioinform 2009;2009:56–60.

29 LePendu P, Iyer SV, Fairon C, et al. Annotation Analysis for Testing Drug Safety Signals using Unstructured Clinical Notes. Journal of Biomedical Semantics. 2012;3. doi:10.1186/2041-1480-3-s1-s5

30 Mikolov T, Sutskever I, Chen K, et al. Distributed Representations of Words and Phrases and their Compositionality. In: Burges CJC, Bottou L, Welling M, et al., eds. Advances in Neural Information Processing Systems 26. Curran Associates, Inc. 2013. 3111–9.

31 McInnes L, Healy J, Melville J. UMAP: Uniform Manifold Approximation and Projection for Dimension Reduction. arXiv [stat.ML]. 2018.http://arxiv.org/abs/1802.03426

32 Jensen PB, Jensen LJ, Brunak S. Mining electronic health records: towards better research applications and clinical care. Nat Rev Genet 2012;13:395–405.

33 Tatonetti NP, Ye PP, Daneshjou R, et al. Data-driven prediction of drug effects and interactions. Sci Transl Med 2012;4:125ra31.

34 Li L, Cheng W-Y, Glicksberg BS, et al. Identification of type 2 diabetes subgroups through topological analysis of patient similarity. Sci Transl Med 2015;7:311ra174.

35 Miotto R, Weng C. Case-based reasoning using electronic health records efficiently identifies eligible patients for clinical trials. J Am Med Inform Assoc 2015;22:e141–50.

36 Boland MR, Parhi P, Li L, et al. Uncovering exposures responsible for birth season--disease effects: a global study. J Am Med Inform Assoc 2018;25:275–88.

37 Xiao C, Choi E, Sun J. Opportunities and challenges in developing deep learning models using electronic health records data: a systematic review. J Am Med Inform Assoc 2018;25:1419–28.

38 Sheikhalishahi S, Miotto R, Dudley JT, et al. Natural Language Processing of Clinical Notes on Chronic Diseases: Systematic Review. JMIR Med Inform 2019;7:e12239.

39 Miotto R, Percha BL, Glicksberg BS, et al. Identifying Acute Low Back Pain Episodes in Primary Care Practice From Clinical Notes: Observational Study. JMIR Med Inform 2020;8:e16878.

40 Landi I, Glicksberg BS, Lee H-C, et al. Deep Representation Learning of Electronic Health Records to Unlock Patient Stratification at Scale. arXiv [q-bio.QM]. 2020.http://arxiv.org/abs/2003.06516

41 Lee J, Liu C, Kim JH, et al. Comparative Effectiveness of Knowledge Graphs-and EHR Data-Based Medical Concept Embedding for Phenotyping. doi:10.1101/2020.07.14.20151274

42 Rajkomar A, Oren E, Chen K, et al. Scalable and accurate deep learning with electronic health records. npj Digital Medicine 2018;1:18.

43 Mullenbach J, Swartz J, Greg McKelvey T, et al. Knowledge Base Completion for Constructing Problem-Oriented Medical Records. arXiv [cs.LG]. 2020.http://arxiv.org/abs/2004.12905

44 Lipton ZC, Kale DC, Elkan C, et al. Learning to Diagnose with LSTM Recurrent Neural Networks. In: ICLR. 2015. 1–18.

45 Miotto R, Li L, Kidd BA, et al. Deep Patient: An Unsupervised Representation to Predict the Future of Patients from the Electronic Health Records. Sci Rep 2016;6:26094.

46 Beaulieu-Jones BK, Greene CS, Pooled Resource Open-Access ALS Clinical Trials Consortium. Semi-supervised learning of the electronic health record for phenotype stratification. J Biomed Inform 2016;64:168–78.

47 Li Y, Rao S, Solares JRA, et al. BEHRT: Transformer for Electronic Health Records. Sci Rep 2020;10:7155.

48 Rasmy L, Xiang Y, Xie Z, et al. Med-BERT: pre-trained contextualized embeddings on large-scale structured electronic health records for disease prediction. arXiv [cs.CL]. 2020.http://arxiv.org/abs/2005.12833

