## Supplementary Material for "Phe2vec: Automated Disease Phenotyping based on Unsupervised Embeddings from Electronic Health Records"

### PheKB Implementation Details

For all PheKB algorithms, we included International Classification of Diseases, 9th revision, (ICD-9) codes for disease diagnoses; Current Procedural Terminology, version 4, (CPT-4) and CPT-Healthcare Common Procedure Coding System (HCPCS) codes for procedures; Logical Observation Identifiers Names and Codes (LOINC) codes and descriptions for lab tests and vital signs. For all non-five digit ICD-9 codes, we used wildcard characters at the end unless otherwise specified (e.g. 314.xx). We obtained medication records by querying each term surrounded by wildcard characters in order to include terms that differ by dosage and administration route (e.g., “%Melipramine%”). All PheKB algorithms were implemented as directed with only the few minor modifications described as follows.

Herpes zoster: we did not implement the antiviral medication minimum dosage threshold as we were not performing association testing in the disease cohorts in the study.

Type 2 diabetes mellitus (T2D): we were not able to distinguish “fasting” from “non-fasting” measurements for glucose lab tests in the Mount Sinai Health System (MSHS) data. Therefore, we considered all records as “non-fasting”. The authors of the algorithm for T2D utilize RxNorm codes for medication supplies, however we sometimes did not have the necessary mappings. Therefore we retrieved some diabetes medical supplies using the same search approach as done for medications. For “Blood-glucose meters and sensors”, we queried: (1) “glucometer”; and (2) “%glucose%” AND (“%meter%  OR “%monitor%” OR “%sensor%”), producing 47 distinct terms. Similarly, for “Insulin syringes”, we queried: “%insulin%” AND (“%syringe%” OR “%inject%” OR “%pen%” OR “%innolet%” OR “%flextouch%” OR “%solostar%” OR “%cart%”), resulting in 356 unique items.

Abdominal aortic aneurysm (AAA): we did not distinguish between case types. Patients having an AAA repair procedure, at least one vascular clinic encounter with diagnosis of ruptured AAA, or at least two vascular clinical encounters with diagnosis of unruptured AAA, were all considered equivalent cases.

Autism: autistic disorder, Asperger’s and PDD-NOS defined by DSM-IV criteria were all considered equivalent cases since they comprise autism spectrum disorder as defined by newly released clinical guidelines in DSM-V.

Multiple Sclerosis: we did not distinguish between case types. Patients with ICD codes for “multiple sclerosis” were considered equivalent to patients with either “demyelinating disease of the central nervous system unspecified”, or “unspecified cause of encephalitis, myelitis, and encephalomyelitis”, or “acute transverse myelitis in demyelinating disease of central nervous system”.

**Supplementary Table 1:** List of data types used in the implementation of PheKB algorithms for each disease included in the study.

| Disease | ICD-9/10 | CPT-4 | Medications | Notes | Labs |
|---|---|---|---|---|---|
| Abdominal aortic aneurysm | ✓ | ✓ | | | |
| Attention deficit hyperactivity disorder | ✓ | | ✓ | | |
| Atrial fibrillation | ✓ | ✓ | | ✓ | |
| Autism | ✓ | | ✓ | ✓ | |
| Crohn’s disease | ✓ | | ✓ | | |
| Dementia | ✓ | | ✓ | | |
| Herpes zoster | ✓ | ✓ | ✓ | | |
| Multiple sclerosis | ✓ | | ✓ | ✓ | |
| Sickle cell disease | ✓ | | | | |
| Type 2 diabetes mellitus | ✓ | | ✓ | | ✓ |

*ICD-9/10: International Classification of Diseases, 9th and 10th revision codes.*
*CPT-4: Current Procedural Terminology, version 4.*

**Supplementary Table 2:** List of all concepts included in the PheKB phenotypes for each disease considered.

| Disease | PheKB Medical Concepts |
|---|---|
| Abdominal aortic aneurysm | **441.3** Abdominal aneurysm, ruptured; **441.4** Abdominal aneurysm without mention of rupture; **CPT** Endovascular repair of infrarenal abdominal aortic aneurysm or dissection using aorto-uniiliac or aorto-unifemoral prosthesis **CPT** Endovascular repair of infrarenal abdominal aortic aneurysm or dissection using modular bifurcated prosthesis **CPT** Endovascular repair of infrarenal abdominal aortic aneurysm or dissection; using unibody bifurcated prosthesis **CPT** Endovascular repair of infrarenal abdominal aortic aneurysm or dissection; using aorto-aortic tube prosthesis **CPT** Open repair of infrarenal aortic aneurysm or dissection, plus repair of associated arterial trauma, following unsuccessful endovascular repair; tube prosthesis **CPT** Open repair of infrarenal aortic aneurysm or dissection, plus repair of associated arterial trauma, following unsuccessful endovascular repair; aorto-bi-iliac prosthesis **CPT** Open repair of infrarenal aortic aneurysm or dissection, plus repair of associated arterial trauma, following unsuccessful endovascular repair; aorto-bifemoral prosthesis **CPT** Direct repair of aneurysm, pseudoaneurysm, or excision (partial or total) and graft insertion, with or without patch graft; for aneurysm, pseudoaneurysm, and associated occlusive disease, abdominal aorta involving visceral vessels (mesenteric, celiac, renal) **CPT** Direct repair of aneurysm, pseudoaneurysm, or excision (partial or total) and graft insertion, with or without patch graft; for aneurysm, pseudoaneurysm, and associated occlusive disease, abdominal aorta **CPT** Direct repair of aneurysm, pseudoaneurysm, or excision (partial or total) and graft insertion, with or without patch graft; for ruptured aneurysm, abdominal aorta **CPT** Direct repair of aneurysm, pseudoaneurysm, or excision (partial or total) and graft insertion, with or without patch graft; for aneurysm, pseudoaneurysm, and associated occlusive disease, hepatic, celiac, renal, or mesenteric artery **CPT** Direct repair of aneurysm, pseudoaneurysm, or excision (partial or total) and graft insertion, with or without patch graft; for ruptured aneurysm, hepatic, celiac, renal, or mesenteric artery **CPT** Direct repair of aneurysm, pseudoaneurysm, or excision (partial or total) and graft insertion, with or without patch graft; for aneurysm, pseudoaneurysm, and associated occlusive disease, abdominal aorta involving iliac vessels (common, hypogastric, external) **CPT** Direct repair of aneurysm, pseudoaneurysm, or excision (partial or total) and graft insertion, with or without patch graft; for aneurysm, pseudoaneurysm, and associated occlusive disease, iliac artery (common, hypogastric, external) **CPT** Direct repair of aneurysm, pseudoaneurysm, or excision (partial or total) and graft insertion, with or without patch graft; for ruptured aneurysm, iliac artery (common, hypogastric, external) |
| Attention deficit hyperactivity disorder | **314** Hyperkinetic syndrome of childhood; **314.0** Attention deficit disorder of childhood; **314.01** Attention deficit disorder with hyperactivity; **314.2** Hyperkinesis with developmental delay; **314.8** Hyperkinetic conduct disorder; Other specified manifestations of hyperkinetic syndrome; **314.9** Unspecified hyperkinetic syndrome; **Med** Imipramine; **Med** Paxil; **Med** Prozac; **Med** Zoloft; **Med** Clonidine; **Med** Carbamazepine; **Med** Clonazepam; **Med** Amphetamine; **Med** Fluoxetine; **Med** Hydroxyzine; **Med** Methylphenidate; **Med** Sertraline; **Med** Paroxetine; **Med** Depakote; **Med** Zyprexa; **Med** Klonopin; **Med** Adderall; **Med** Atarax; **Med** Trazodone; **Med** Sarafem; Med Focalin; **Med** Concerta; **Med** Risperdal; **Med** Methylin; **Med** Vistaril; **Med** Tegretol; **Med** Wellbutrin; **Med** Dexedrine; **Med** Dexmethylphenidate; **Med** Desoxyn; **Med** Ritalin; **Med** Metadate; **Med** Lithobid; **Med** Strattera; **Med** Risperidone; **Med** Pexeva; **Med** Olanzapine; **Med** Pemoline; **Med** Guanfacine; **Med** Equetro; **Med** Tofranil; **Med** Zyban; **Med** Daytrana; **Med** Lisdexamfetamine; **Med** Vyvanse; **Med** Aplenzin; **Med** Atomoxetine; **Med** Lithium; **Med** Oleptro; **Med** Brisdelle |

| | |
|---|---|
| Atrial fibrillation | **427.31** Atrial fibrillation; **427.32** Atrial flutter; |
| Autism | **290.00** Autistic disorder, current or activate state; **290.01** Autistic disorder, residual state; **299.80** Other specified pervasive developmental disorders, current or active **299.81** Other specified pervasive developmental disorders, residual state **299.90** Unspecified pervasive developmental disorder, current or active state **299.91** Unspecified pervasive developmental disorder, residual state |
| Crohn's disease | **555** Regional enteritis; **555.1** Regional enteritis of large intestine; **555.2** Regional enteritis of small intestine with large intestine; **555.9** Regional enteritis of unspecified site; **Med** Adalimumab; **Med** Asacol; **Med** Azathioprine; **Med** Azulfidine; **Med** Balsalazide; **Med** Budesonide; **Med** Canasa; **Med** Certolizumab pegol; **Med** Cimzia; **Med** Ciprofloxacin; **Med** Colazal; **Med** Humira; **Med** Infliximab; **Med** Levofloxacin; **Med** Lialda; **Med** Mercaptopurine; **Med** Mesalamine; **Med** Metronidazole; **Med** Natalizumab; **Med** Pentasa; **Med** Prednisone; **Med** Purinethol; **Med** Remicade; **Med** Rifaximin; **Med** Rowasa; **Med** Sulfasalazine; **Med** Tysabri |
| Dementia | **290.0** Senile dementia, uncomplicated; **290.10** Presenile dementia, uncomplicated; **290.11** Presenile dementia with delirium; **290.12** Presenile dementia with delusional features; **290.13** Presenile dementia with depressive features; **290.20** Senile dementia with delusional features; **290.21** Senile dementia with depressive features; **290.3** Senile dementia with delirium; **290.40** Vascular dementia, uncomplicated; **290.41** Vascular dementia, with delirium; **290.42** Vascular dementia, with delusions; **290.43** Vascular dementia, with depressed mood; **291.0** Alcohol withdrawal delirium; **291.1** Alcohol-induced persisting amnestic disorder; **291.2** Alcohol-induced persisting dementia; **292.82** Drug-induced persisting dementia; **294.10** Dementia in conditions classified elsewhere without behavioral disturbance; **294.11** Dementia in conditions classified elsewhere with behavioral disturbance; **294.8** Other persistent mental disorders due to conditions classified elsewhere; **331.0** Alzheimer's disease; **331.11** Pick's disease; **331.19** Other frontotemporal dementia; **331.82** Dementia with lewy bodies; **Med** Aricept; **Med** Exelon; **Med** Namenda; **Med** Razadyne; |
| Herpes zoster | **52** Chickenpox; **52.0** Postvaricella encephalitis; **52.1** Varicella (hemorrhagic) pneumonitis; **52.2** Postvaricella myelitis; **52.7**Chickenpox with other specified complications; **52.8** Chickenpox with unspecified complication; **52.9** Varicella without mention of complication; **53** Herpes zoster; **53.0** Herpes zoster with meningitis; **53.1** Herpes zoster with other nervous system complications; **53.1** Herpes zoster with unspecified nervous system complication; **53.11** Geniculate herpes zoster; **53.12** Postherpetic trigeminal neuralgia; **53.13** Postherpetic polyneuropathy; **53.14** Herpes zoster myelitis; **53.19** Herpes zoster with other nervous system complications; **53.2** Herpes zoster with ophthalmic complications; **53.2** Herpes zoster dermatitis of eyelid; **53.21** Herpes zoster keratoconjunctivitis; **53.22** Herpes zoster iridocyclitis; **53.29** Herpes zoster with other ophthalmic complications; **53.7** Herpes zoster with other specified complications; **53.71** Otitis externa due to herpes zoster; **53.79** Herpes zoster with other specified complications; **53.8** Herpes zoster with unspecified complication; **53.9** Herpes zoster without mention of complication |
| Multiple sclerosis | **323.9** Unspecified causes of encephalitis, myelitis, and encephalomyelitis; **341.20** Acute (transverse) myelitis nos; **341.21** Acute (transverse) myelitis in conditions classified elsewhere; **341.9** Demyelinating disease of central nervous system, unspecified; **Med** Avonex; **Med** Copaxone; **Med** Betaseron; **Med** Interferon beta-1b; **Med** Interferon beta-1a; **Med** Rebif; **Med** Rebif titration pack; **Med** Glatiramer acetate; **Med** Peginterferon beta 1a; |
| Sickle cell disease | **282.41** Sickle-cell thalassemia without crisis; **282.42** Sickle-cell thalassemia with crisis; **282.60** Sickle-cell disease, unspecified; **282.61** Hb-ss disease without crisis; **282.62** Hb-ss disease with crisis; **282.63** Sickle-cell/hb-c disease without |

| | |
|---|---|
| | crisis; **282.64** Sickle-cell/hb-c disease with crisis; **282.68** Other sickle-cell disease without crisis; **282.69** Other sickle-cell disease with crisis |
| Type 2 diabetes mellitus | **250** Diabetes mellitus without mention of complication; **250** Diabetes mellitus without mention of complication, type ii or unspecified type, not stated as uncontrolled; **250.02** Diabetes mellitus without mention of complication, type ii or unspecified type, uncontrolled; **250.1** Diabetes with ketoacidosis; **250.1** Diabetes with ketoacidosis, type ii or unspecified type, not stated as uncontrolled; **250.12** Diabetes with ketoacidosis, type ii or unspecified type, uncontrolled; **250.2** Diabetes mellitus with hyperosmolarity; **250.2** Diabetes with hyperosmolarity, type ii or unspecified type, not stated as uncontrolled; **250.22** Diabetes with hyperosmolarity, type ii or unspecified type, uncontrolled; **250.3** Diabetes with other coma; **250.3** Diabetes with other coma, type ii or unspecified type, not stated as uncontrolled; **250.32** Diabetes with other coma, type ii or unspecified type, uncontrolled; **250.4** Diabetes with renal manifestations; **250.4** Diabetes with renal manifestations, type ii or unspecified type, not stated as uncontrolled; **250.42** Diabetes with renal manifestations, type ii or unspecified type, uncontrolled; **250.5** Diabetes with ophthalmic manifestations; **250.5** Diabetes with ophthalmic manifestations, type ii or unspecified type, not stated as uncontrolled; **250.52** Diabetes with ophthalmic manifestations, type ii or unspecified type, uncontrolled; **250.6** Diabetes with neurological manifestations; **250.6** Diabetes with neurological manifestations, type ii or unspecified type, not stated as uncontrolled; **250.62** Diabetes with neurological manifestations, type ii or unspecified type, uncontrolled; 250.7 Diabetes with peripheral circulatory disorders; **250.7** Diabetes with peripheral circulatory disorders, type ii or unspecified type, not stated as uncontrolled; **250.72** Diabetes with peripheral circulatory disorders, type ii or unspecified type, uncontrolled; **250.8** Diabetes with other specified manifestations; **250.8** Diabetes with other specified manifestations, type ii or unspecified type, not stated as uncontrolled; 2**50.82** Diabetes with other specified manifestations, type ii or unspecified type, uncontrolled; **250.9** Diabetes with unspecified complication; **250.9** Diabetes with unspecified complication, type ii or unspecified type, not stated as uncontrolled; **250.92** Diabetes with unspecified complication, type ii or unspecified type, uncontrolled **Med** Acarbose; **Med** Chlorpropamide; **Med** Exenatide; **Med** Glimepiride; **Med** Glipizide; **CPT** Glucose; **CPT** Glucose tolerance; **Med** Glyburide; **CPT** Hemoglobin A1c; **Med** Metformin; **Med** Miglitol; **Med** Nateglinide; **Med** Pioglitazone; **Med** Repaglinide; **Med** Rosiglitazone; **Med**Sitagliptin |

**Supplementary Table 3:** List of seed medical concepts used to define phenotypes with Phe2vec for all diseases considered in the study. The seed selected is the ICD-9 code that most closely represents the disease.

| Disease | ICD-9 Seed Concept |
|---|---|
| Abdominal aortic aneurysm | 441.4, Abdominal aneurysm without mention of rupture |
| Attention deficit hyperactivity disorder | 314.01, Attention deficit disorder with hyperactivity |
| Atrial fibrillation | 427.31, Atrial fibrillation |
| Autism | 299.00, Autistic disorder, current or active state |
| Crohn's disease | 555.9, Regional enteritis of unspecified site |
| Dementia | 294.20, Dementia, unspecified, without behavioral disturbance |
| Herpes zoster | 053.9, Herpes zoster without mention of complication |
| Multiple sclerosis | 340, Multiple sclerosis |
| Sickle cell disease | 282.60, Sickle-cell disease, unspecified |
| Type 2 diabetes mellitus | 250.00, Diabetes mellitus without mention of complication, type ii or unspecified type, not stated as uncontrolled |
| Lyme disease | 088.81, Lyme disease |

*ICD-9: International Classification of Diseases, 9th revision*

**Supplementary Figure 1:** Uniform manifold approximation and projection (UMAP) visualization of the EHR-based phenotype space generated by Phe2vec with word2vec embeddings for (A) Abdominal aortic aneurysm; (B) Attention deficit hyperactivity disorder; (C) Atrial fibrillation; (D) Autism; (E) Crohn's disease; (F) Multiple sclerosis; (G) Sickle cell disease; (H) Herpes zoster. Seed concepts are colored in black, while concepts in the phenotypes are colored in purple.

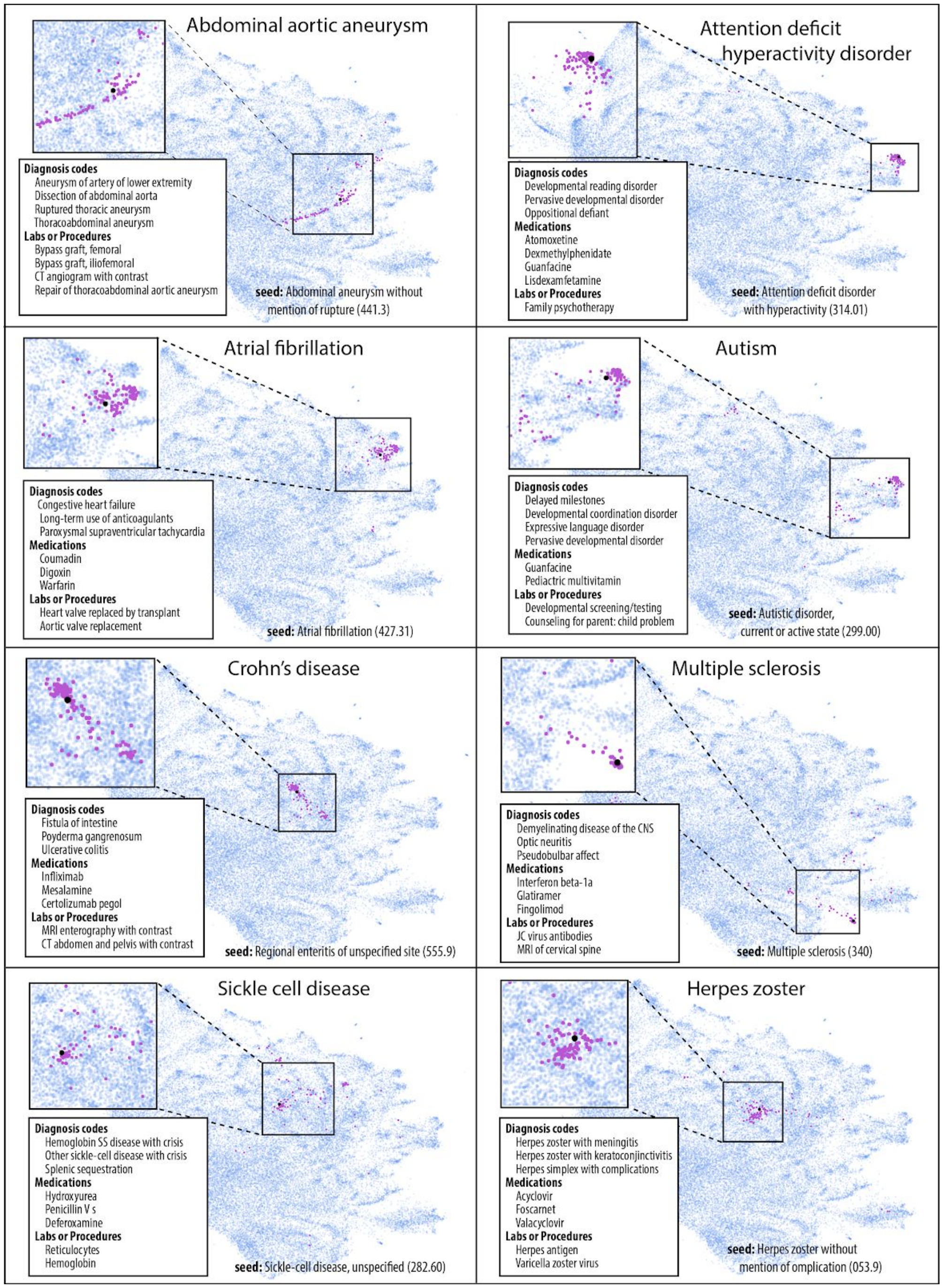

**Supplementary Table 4:** Results of cohort selection per disease obtained by Phe2vec, with GloVe embeddings, where PheKB cohorts are considered as gold standard.

| Disease | Patients | F-score | R-precision | AUC-PR |
|---|---|---|---|---|
| Abdominal aortic aneurysm | 1,982 | 0.58 | 0.60 | 0.69 |
| Attention deficit hyperactivity disorder | 7,778 | 0.67 | 0.68 | 0.79 |
| Atrial fibrillation | 39,568 | 0.50 | 0.47 | 0.51 |
| Autism | 1,279 | 0.49 | 0.45 | 0.48 |
| Crohn's disease | 6,207 | 0.67 | 0.61 | 0.72 |
| Dementia | 15,406 | 0.46 | 0.52 | 0.51 |
| Herpes zoster | 1,618 | 0.42 | 0.33 | 0.48 |
| Multiple sclerosis | 4,532 | 0.78 | 0.79 | 0.82 |
| Sickle cell disease | 949 | 0.53 | 0.51 | 0.56 |
| Type 2 diabetes mellitus | 59,233 | 0.59 | 0.52 | 0.60 |

**Supplementary Table 5:** Results of cohort selection per disease obtained by Phe2vec, with FastText embeddings, where PheKB cohorts are considered as gold standard.

| Disease | Patients | F-score | R-precision | AUC-PR |
|---|---|---|---|---|
| Abdominal aortic aneurysm | 1,982 | 0.63 | 0.57 | 0.69 |
| Attention deficit hyperactivity disorder | 7,778 | 0.76 | 0.66 | 0.78 |
| Atrial fibrillation | 39,568 | 0.51 | 0.49 | 0.53 |
| Autism | 1,279 | 0.50 | 0.48 | 0.54 |
| Crohn's disease | 6,207 | 0.72 | 0.61 | 0.74 |
| Dementia | 15,406 | 0.45 | 0.49 | 0.47 |
| Herpes zoster | 1,618 | 0.38 | 0.31 | 0.56 |
| Multiple sclerosis | 4,532 | 0.83 | 0.79 | 0.86 |
| Sickle cell disease | 949 | 0.55 | 0.62 | 0.58 |
| Type 2 diabetes mellitus | 59,233 | 0.61 | 0.53 | 0.71 |

**Supplementary Table 6:** Results of cohort selection per disease obtained by BoCon, where PheKB cohorts are considered as gold standard.

| Disease | Patients | F-score | R-precision | AUC-PR |
|---|---|---|---|---|
| Abdominal aortic aneurysm | 1,982 | 0.56 | 0.57 | 0.62 |
| Attention deficit hyperactivity disorder | 7,778 | 0.65 | 0.56 | 0.64 |
| Atrial fibrillation | 39,568 | 0.39 | 0.26 | 0.42 |
| Autism | 1,279 | 0.53 | 0.48 | 0.54 |
| Crohn's disease | 6,207 | 0.52 | 0.39 | 0.65 |
| Dementia | 15,406 | 0.42 | 0.32 | 0.47 |
| Herpes zoster | 1,618 | 0.25 | 0.19 | 0.28 |
| Multiple sclerosis | 4,532 | 0.84 | 0.73 | 0.81 |
| Sickle cell disease | 949 | 0.63 | 0.34 | 0.65 |
| Type 2 diabetes mellitus | 59,233 | 0.54 | 0.51 | 0.55 |

**Supplementary Figure 2:** Inter-rater reliability between each pair of raters evaluated using percent agreement. The value was calculated from the independent review of presumed cases by each rater, prior to establishing a consensus on those for which raters disagreed on disease status.

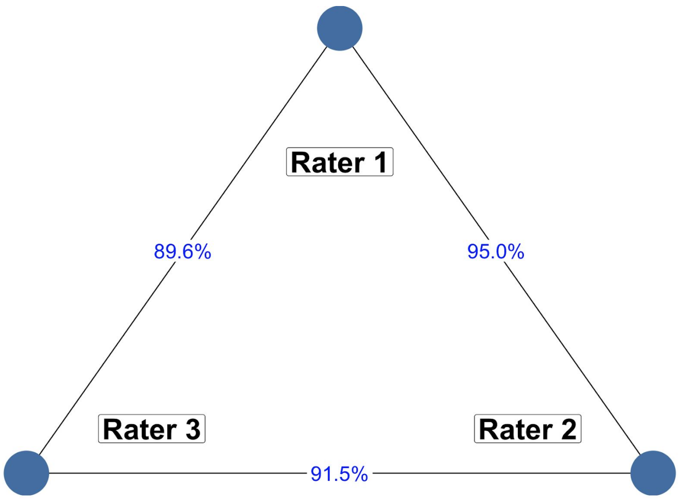
